# Frontotemporal Dementia Subtyping using Machine Learning, Multivariate Statistics, and Neuroimaging

**DOI:** 10.1101/2024.06.17.24308959

**Authors:** Amelie Metz, Yashar Zeighami, Simon Ducharme, Sylvia Villeneuve, Mahsa Dadar

**Affiliations:** Integrated Program in Neuroscience, McGill University, Montreal, H3A 2B4, Canada; Douglas Mental Health University Institute, Montreal, H4H 1R3, Canada; McConnell Brain Imaging Centre, Montreal Neurological Institute, McGill University, Montreal, H3A 2B4, Canada; Department of Psychiatry, McGill University, Montreal, H3A 1A1, Canada

**Keywords:** frontotemporal dementia, primary progressive aphasia, multivariate statistics, machine learning, neuroimaging, diagnostic accuracy

## Abstract

Frontotemporal Dementia (FTD) is a prevalent form of early-onset dementia characterized by progressive neurodegeneration. It encompasses a group of heterogeneous disorders, including behavioral variant frontotemporal dementia (bvFTD), nonfluent variant primary progressive aphasia (nfvPPA), and semantic variant primary progressive aphasia (svPPA). Due to disease heterogeneity and overlapping symptoms, diagnosis of FTD and its subtypes still poses a challenge. Magnetic-resonance imaging (MRI) is commonly used to support the diagnosis of FTD. Using machine learning and multivariate statistics, we tested whether brain atrophy patterns are associated with severity of cognitive impairment, whether this relationship differs between the phenotypic subtypes, and whether we could use these brain patterns to classify patients according to their FTD variant.

A total of 136 patients (70 bvFTD, 36 svPPA, 30 nfvPPA) from the frontotemporal lobar degeneration neuroimaging initiative (FTLDNI) database underwent brain MRI and clinical and neuropsychological examination. Deformation-based morphometry (DBM), which offers increased sensitivity to subtle local differences in structural image contrasts was used to estimate regional cortical and subcortical atrophy. Atlas-based associations between DBM values and performance across different cognitive tests were assessed using partial least squares (PLS). We then applied linear regression models to discern the group differences regarding the relationship between atrophy and cognitive decline in the three FTD phenotypes. Lastly, we assessed whether the combination of neural and behavioral patterns in the latent variables identified in the PLS analysis could be used as features in a machine-learning model to predict FTD subtypes in patients.

Results revealed four significant latent variables that combined accounted for 86% of the shared covariance between cognitive and brain atrophy measures. PLS-based atrophy and behavioral patterns predicted the FTD phenotypes with a cross-validated accuracy of 89.12%, with high specificity (91.46-97.15%) and sensitivity (84.19-93.56%). When using only MRI measures and two behavioral tests in the PLS and classification algorithm, ensuring clinical feasibility, our model was similarly precise (83.62%, specificity 86.38-93.51%, sensitivity 76.17-87.50%). Here, including only atrophy or behavior patterns in the analysis led to prediction accuracies of 69.76% and 76.38%, respectively, highlighting the increased value of combining MRI and clinical measures in subtype classification.

We demonstrate that the combination of brain atrophy and clinical characteristics, and multivariate statistical methods can serve as an imaging biomarker for early disease phenotyping in FTD, whereby inclusion of DBM measures adds to the classification precision in the absence of extensive clinical testing.

## Introduction

Frontotemporal Dementia (FTD) is one of the most common forms of early-onset dementia. It is characterized by atrophy and gliosis in the frontal and temporal lobes of the brain^1^ as well as neuropathological abnormalities in the form of hyperphosphorylated protein accumulations, typically composed of either tau or TDP-43^2,3^. Clinically, it encompasses a group of heterogeneous neuropathological disorders causing a wide spectrum of symptoms, including changes in behavior, language, executive control, and motor symptoms^4,5^. The core FTD spectrum syndromes are behavioral variant frontotemporal dementia (bvFTD), nonfluent variant primary progressive aphasia (nfvPPA), and semantic variant primary progressive aphasia (svPPA)^6^. Patients with bvFTD initially present with abnormal behavior, changes in personality and emotion, and reduced executive control and social cognition. This includes symptoms like disinhibition, compulsions, dietary changes, apathy, or lack of empathy^7^. In primary progressive aphasias, cognitive deficits predominantly manifest themselves in the language domain. About 5 to 7 years after symptom onset, as pathology spreads,^5^ patients develop additional behavioral symptoms of bvFTD. Patients with svPPA show progressive impairments in conceptual knowledge, word retrieval, and single-word comprehension whereas nfvPPA is defined by effortful speech in combination with motor speech apraxia and agrammatism^8^.

Given the heterogeneity and considerable overlap of clinical symptoms and pathology of FTD with other disorders, the diagnosis of FTD still poses a significant challenge to clinicians^9^. To date, no validated FTD-specific plasma, cerebrospinal fluid or positron emission tomography biomarkers have been identified. Structural magnetic resonance imaging (MRI) is therefore commonly used in clinical practice to confirm a diagnosis of FTD^10^. However, MRI is currently insufficiently sensitive to detect subtle neuronal loss in the early stages of the disease^11^, leading to delayed or incorrect diagnoses. This leads to delayed and inappropriate treatment and increased distress for patients and caretakers. The development of biomarkers for the detection and diagnosis of FTD at symptom onset is critical to ensure optimal care for patients as well as to accurately inform critical trials. As methods of morphometric analysis and the use of multivariate statistics and machine learning methods are advancing, it is becoming increasingly feasible to combine MRI-based features with these techniques to improve early detection and diagnosis.

Previous MRI studies on FTD have largely focused on assessing gray matter atrophy as measured by cortical thickness or voxel-based morphometry^10^. The findings of these studies corresponded to evidence from post-mortem studies in that they demonstrated specific patterns of atrophy in frontal and temporal cortices on a group level^12,13^. BvFTD has been related to atrophy in fronto-insular cortices as well as basal ganglia^14–17^. SvPPA has been associated with left anterior temporal pole and hippocampal atrophy^18–21^. In nfvPPA, atrophy seems to be most prevalent in the left inferior frontal gyrus, specifically involving Broca’s area^12,22–25^. Another method to estimate atrophy patterns in MRI images is deformation-based morphometry (DBM). Using DBM has considerable advantages over other estimations of cortical atrophy. While voxel-based morphometry and cortical thickness estimation depend on automated tissue segmentation, occasionally leading to erroneous tissue classification and thereby to incorrect calculation of gray matter volume^26^, DBM does not directly rely on image contrast to represent tissue alterations. DBM is also more sensitive to subtle differences as it matches images locally (i.e. at voxel level) based on similarities in contrast and intensities^27,27,28^. This makes DBM a potential candidate for diagnostic purposes, given the need for improved diagnostic biomarkers for FTD and particularly for the categorization of patients according to FTD variants. However, only few studies have applied DBM in the context of FTD. For instance, studies focusing on bvFTD identified the insula and anterior and medial temporal regions as epicenters of brain atrophy^29,30^. In svPPA, the left medial temporal lobe and perirhinal cortex have been implicated^31^. Cardenas *et al.*^32^ studied DBM in FTD patients and found atrophy in the frontal lobes and anterior temporal lobes as well as in the thalamus, pons, and superior and inferior colliculi. While these findings broadly align with the conclusions of other MRI studies, they also highlight the utility of DBM in identifying small changes in gray matter structure, especially in subcortical areas^33^.

Previous attempts at an automated classification of FTD patients based on structural MRI in combination with machine learning techniques have achieved high accuracy (80-90%) in distinguishing patients from control groups^10^. However, few cases have implemented a multiclass approach^34–40^. As binary classifiers necessitate the exclusion of all but two potential clinical labels, multiclass methods offer greater value from a clinical standpoint. Additionally, FTD subtypes represent a spectrum, making it challenging for a binary variable to fully encompass their nuances. Thus, we opted for a multiclass method here. Furthermore, many studies grouped FTD clinical variants together in their analysis. Considering their heterogeneity in terms of behavioral and neurodegenerative features, early detection of specific FTD syndromes is highly relevant to determine appropriate treatments. While studies achieved high accuracies^22,41^, only two studies classified each FTD subtype against a group of all others and patients with Alzheimer’s Disease. Notably, Tahmasian et al. (2016)^42^ achieved high specificity (97.5% and 94.2%) but very poor sensitivity (50% and 0%). Kim et al. (2019)^43^ used cortical thickness measures in a hierarchical classification scheme to classify FTD subtypes which resulted in an accuracy of 75.8%. Improved implementation of machine learning is therefore crucial for enhancing early detection of FTD variants.

The present study took advantage of the sensitivity of DBM measurements to capture the relationship between brain atrophy patterns and disease-related clinical measures in the three main variants of FTD. Modeling the relationship between brain atrophy and cognitive decline individually in three phenotypic variants (bvFTD, svPPA, nfvPPA) might make it possible to disambiguate the different domains of disease within the FTD population and their link with brain morphometric measures. Here, we used a multivariate method to relate the different cognitive symptoms of FTD variants to system-wide atrophy patterns. We analyzed data from 136 FTD patients who were diagnosed with either bvFTD, svPPA, or nfvPPA from the frontotemporal lobar degeneration neuroimaging initiative (FTLDNI) database. We used DBM as a measure of structural brain alterations, in combination with partial least squares (PLS)^44–46^ to quantify the magnitude and pattern of volume change in different FTD variants as compared to each other and to identify the cortical and subcortical structures most sensitive to change. Using the resulting patterns that maximally explain the covariance between FTD subtypes, we were able to predict FTD variant diagnosis in our cohort.

In concordance with previous neuroimaging and ex vivo studies, we expected pathological changes in fronto- and temporocortical regions as well as subcortical structures. The main aim of this study was to enhance the understanding of the disease mechanisms underlying FTD and to provide potential early imaging biomarkers for disease severity assessment and phenotyping.

## Materials and methods

### Participants

Data analyzed in this study includes participants from the FTLDNI that had T1-weighted MR images. The FTLDNI was founded through the National Institute of Aging and started in 2010 (https://memory.ucsf.edu/research/studies/nifd). The primary goals of FTLDNI are to identify neuroimaging modalities and methods of analysis for tracking frontotemporal lobar degeneration and to assess the value of imaging versus other biomarkers in diagnostic roles. Baseline and follow-up data from 136 FTLDNI participants were included in this study. Data was accessed and downloaded through the LONI platform in July 2023. We included patients with bvFTD (*n*(baseline) = 70, *n*(followup) = 38), svPPA (*n(baseline)* = 36, *n*(followup) = 24), and nfPPA (*n(baseline)* = 30, *n*(followup) = 15). The inclusion criteria for FTD patients were diagnosis of possible or probable FTD according to the FTD consortium criteria^7^. All subjects provided informed consent and the protocol was approved by the institutional review boards at all sites.

### Clinical assessment

All participants were assessed at the initial visit for clinical characteristics (motor, non-motor, and neuropsychological performance) by site investigators. The neuropsychological assessment included Mini-Mental State Examination (MMSE) and Clinical Dementia Rating Scale (CDR) as measures of global cognition, forward digit span and California Verbal Learning Test (CVLT, items recalled correctly after four learning trials, items recalled correctly after 30 second delay, items recalled correctly after 10-minute delay, and word recognition) as measures of verbal memory and learning, Modified Trail Making Test (MTMT, time and correct lines) and backward digit span as measures of executive function, as well as Verbal Fluency (phonological and semantic), Boston Naming Test (BNT), and Peabody Picture Vocabulary Test (PPVT) as measures of language ability.

### Structural MRI acquisition and processing

For details on MRI acquisition protocols and scanner information, please refer to https://cind.ucsf.edu/research/grants/frontotemporal-lobar-degeneration-neuroimaging-initiative-0.

T1w scans for each participant were pre-processed through our standard pipeline including noise reduction^47^, intensity inhomogeneity correction^48^, and intensity normalization into the range [0-100]. The pre-processed images were then linearly (9 parameters: 3 translation, 3 rotation, and 3 scaling)^49^ and nonlinearly^50^ registered to the MNI-ICBM152-2009c average^51^. The quality of the linear registrations was visually verified by an experienced rater (author M.D.), blinded to the diagnostic group. Only 7 scans did not pass this quality control step and were discarded.

### DBM values

DBM analysis was performed using MNI MINC tools^29^. The principle of DBM is to warp each individual scan to a common template through a non-linear deformation, where local shape differences between the two images (i.e., the subject’s T1w image and the template) are encoded in the deformations. The local deformation obtained from the non-linear transformations can then be used as a measure of tissue expansion or atrophy by estimating the determinant of the Jacobian for each transform. Local contractions can be interpreted as shrinkage of tissue (atrophy) and local expansions are often related to ventricular or sulci enlargement. DBM was used to assess regional volumetric differences whereby DBM values were calculated based on 102 regions from CerebrA atlas^52^.

## Statistical analysis

### Demographics and cognitive scores

All statistical analyses were performed using MATLAB version R2022a. One-way ANOVAs were conducted to compare demographic and cognitive variables at baseline, followed by independent t-tests with Tukey HSD correction for multiple comparisons. Categorical variables (i.e., sex) were analyzed using chi-square analyses. Results are expressed as mean±standard deviation and [median]. A p-value of *p* <0.05 was considered statistically significant.

### Partial least squares regression

PLS analysis was used to relate behavioral and brain atrophy patterns^44,46^. The goal of the analysis is to identify combinations of cognitive scores and brain patterns that optimally covary with each other. PLS is a multivariate technique used to establish relationships between two sets of variables. This approach can be used to identify weighted linear combinations of variables that exhibit a high degree of covariation^45^. The resulting linear combinations of these variables can be construed as atrophy networks and their corresponding clinical manifestations.

We followed the approach described in Zeighami et al.^53^ (Fig.1). Behavioral and brain data were represented as two matrices X and Y, with X representing cognitive scores in 12 columns, while Y representing brain measure across 102 regions using the CerebrA atlas. The matrices were standardized through z-scoring and a correlation matrix (**X**□**Y**) was computed. The correlation matrix was then subjected to singular value decomposition (SVD^54^).

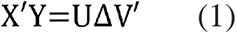

**Figure 1.**
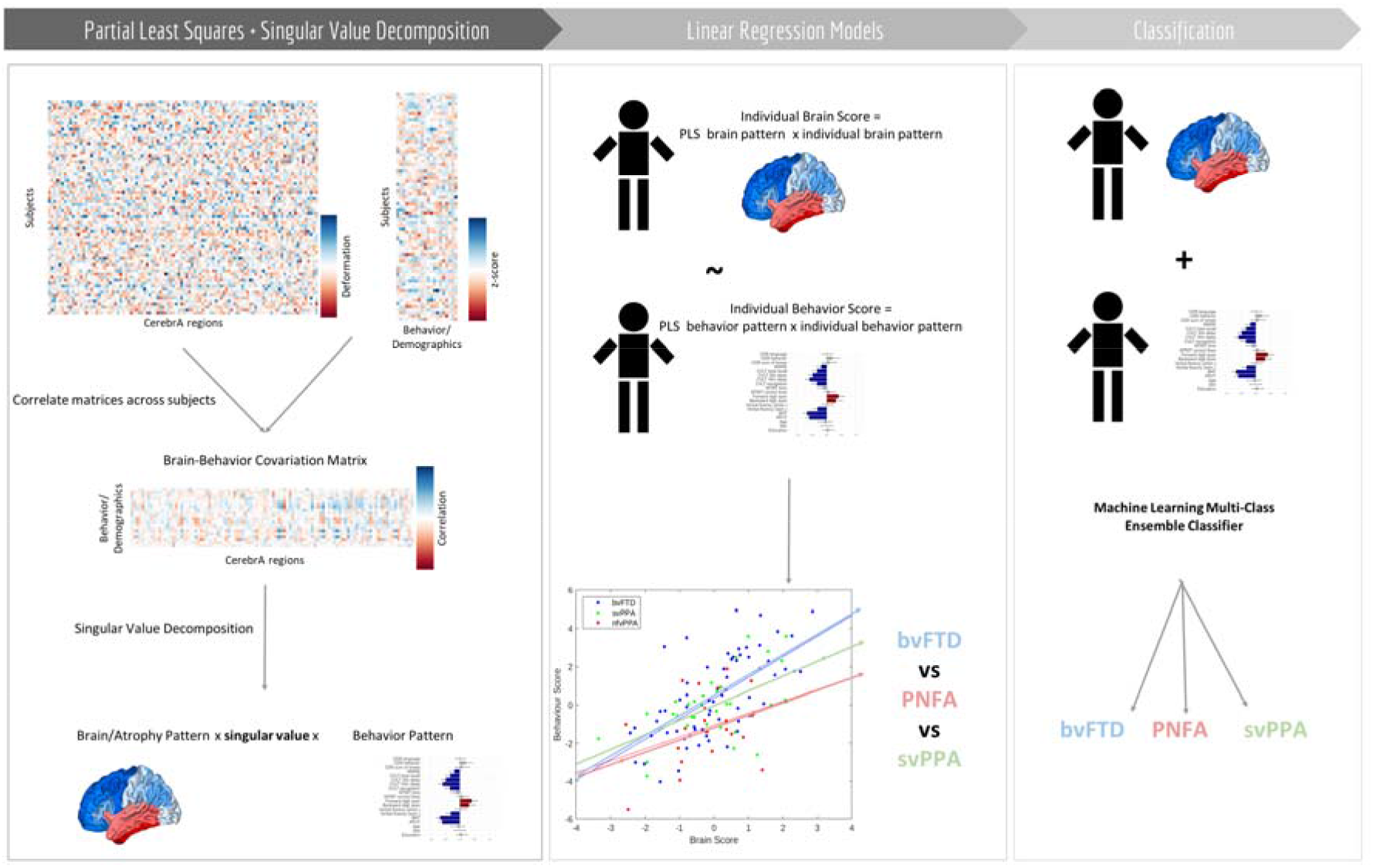
Partial Least Squares (PLS) analysis flowchart. Z-scored matrices for DBM data and behavioral/demographic data were combined into a single brain * behavior covariation matrix. Consequently, we applied single value decomposition to the resulting matrix, yielding orthogonal latent variables (LV). Each LV represented atrophy patterns linked to clinical characteristics, with their associated singular value reflecting the covariance between atrophy and behavior (for detailed explanation see Zeighami et al^53^). We then applied linear regression models to discern the differences between the diagnostic FTD groups in terms of behavior and brain patterns in each LV. Lastly, we used the individual brain and behavior scores of each patient as features in a machine learning classifier (employing an ensemble of discriminant learners with bagging aggregation method) to predict the FTD subtype of each subject both in the baseline and follow-up visit data.

The decomposition process results in a collection of orthogonal latent variables (LVs), with U and V constituting the matrices of left and right singular vectors, while Δ is a diagonal matrix containing the singular values. The covariance explained by each latent variable is used as its effect size.

The statistical significance of each LV was evaluated using permutation tests. The rows of matrix X were randomly permuted (repeated 500 times), and the behavior-brain correlation matrix was recomputed. The permuted correlation matrices were subjected to SVD as before, generating the null distribution for the covariance explained across LVs, which was then used to calculate the *P*-value.

The contribution of individual variables was assessed through bootstrap resampling. By random sampling with replacement of participants (repeated 500 times). As a result, a sampling distribution was generated for each individual weight within the singular vectors. A "bootstrap ratio" was computed for each CerebrA region, representing the ratio of its singular vector weight to its standard error estimated through bootstrapping and used to identify voxels that make substantial contributions to the atrophy patterns^55^. Bootstrap ratio maps were thresholded using the 95% confidence interval criterion.

### Group differences between FTD variants

To gauge the extent to which the data driven PLS patterns differ between the FTD variants, patient-specific scores were computed. Specifically, the brain and behavioural patterns for latent variables were projected onto individual patients’ data, producing scalar atrophy scores and behavioral scores for each subject. These scores are akin to principal component scores or factor scores:

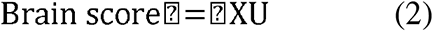

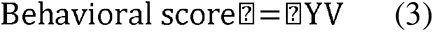

We then used the following Linear Regression Model to assess if the relationship between atrophy scores and behavioral scores differs between the three phenotypic FTD groups:

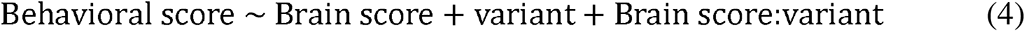

The variable of interest was the interaction term, Brain score:variant, which indicates the slope differences between the three FTD subtypes and reflects the contribution of cortical atrophy to cognitive performance in each diagnostic group.

### Predictive capacity of the model

We assessed whether the combination of brain and behavioral patterns identified in the PLS analysis could be used as features to predict FTD subtypes in patients. To this end, we employed a machine learning multi-class ensemble classifier, using an ensemble of discriminant learners (bagging aggregation method). A 10-fold cross-validation scheme was performed on 100 randomized train and test splits to assess the performance of the classifier. We used the combination of neural and behavioral patterns in the latent variables identified in the PLS analysis as features to train the model. The model then predicted clinical diagnoses (i.e. FTD variants) in the test data. Performance of the model was evaluated in terms of prediction accuracy for each cross-validation fold (mean and standard deviation) by comparing the predicted diagnostic group with the clinician diagnosis (‘gold standard’) in the cross-validated test subset. We also examined the sensitivity and specificity of our model for each FTD subtype. Note that the FTD variant information was not included in any previous steps (i.e. the PLS analysis), to avoid leakage of information in the variant classification task. As our model included measures of disease severity, we also tested the precision of the predictions in a severity-matched sample, based on CDR scores (only including participants with CDR scores below 1.5), to ensure that the model does not solely rely on group differences in disease severity.

Finally, we validated the stability of our model by projecting brain and behavior patterns of the LVs onto the longitudinal data to predict patients’ FTD subtype diagnosis. Using the same machine learning multi-class ensemble classifier as before, we utilized the baseline data as the training set and tested the performance of the model on the longitudinal data as the test set. Note that prediction for the longitudinal timepoints was also performed within the same cross-validation scheme, and no data from baseline visits of the same participants were used in the training folds for the longitudinal predictions. We converted follow-up test and DBM scores into z-scores based on the baseline data, by matching each score to the respective baseline z-score. This ensured that follow-up values were comparable to the baseline (cross-sectional) dataset. We again measured the prediction accuracy, as well as sensitivity and specificity, of the model in identifying the diagnosed FTD variant of the patient.

To ensure the clinical utility of our diagnostic approach in terms of time and necessary neuropsychological test batteries, we repeated the PLS analysis and prediction while including minimal variables that can be completed in under an hour, i.e. only using T1w MRI derived DBM values for the brain patterns (5-10 minutes), and CDR (box score, language score, and behavior score, ∼30minutes) and BNT (5-15minutes) for the behavior score. As the full battery of tests included in our PLS analysis takes over 2 hours to administer, a minimal assessment including CDR and BNT is more feasible and still informative for clinicians.

### Longitudinal changes

To determine the longitudinal change of the brain and behavior patterns resulting from the PLS analysis, we compared the scores of each patient with a baseline and follow-up visit (*n* = 32) using pairwise t-tests. Lastly, we calculated the yearly rate of change in behavior and brain scores for each FTD syndrome to investigate whether the progression of clinical symptoms and atrophy patterns differs between diagnostic groups. We then used unpaired t-tests to compare the rates of change between the FTD variants.

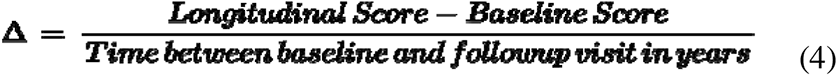

This analysis was repeated for the brain and behavior scores of each LV separately. For longitudinal measurements, we selected the time point for each subject that was closest to a one-year follow-up after the baseline assessment. The mean time between baseline and follow-up visits was 1.03 years (*sd* = 0.44) with a range of 0.40 - 3.57 years.

### Data availability

FTLDNI MRI and clinical measures are available through https://ida.loni.usc.edu/login.jsp. Derived data supporting the findings of this study, including individual PLS scores, are available from the corresponding author on request.

## Results

### Demographic and clinical characteristics

Table 1 compares demographic variables and cognitive test scores between bvFTD, svPPA, and nfvPPA patients at baseline. Significant differences were observed in age distribution, with nfvPPA subjects being on average older compared to other groups (bvFTD: p< 001; svPPA: p<.007). Clinical measures also showed notable distinctions (*Supplementary Table* 1).

**Table 1.** Baseline demographic and cognitive characteristics in FTD subtypes. . Values expressed as mean (standard deviation). Asterisks indicate significant group differences based on one-way ANOVA or chi-square analysis comparing the groups. bvFTD = behavioral variant FTD; svPPA = semantic variant primary progressive aphasia; nfvPPA = nonfluent variant primary progressive aphasia.

|  | bvFTD | svPPA | nfvPPA | p-value |
| --- | --- | --- | --- | --- |
| **Demographics** |  |  |  |  |
| *Number of subjects (total n=136)* | 70 | 36 | 30 |  |
| *Age (years, mean (SD))* | 61.47 (6.39) | 63 (6.30) | 68.10 (7.90) | <.001* |
| *Sex (M:F, %M)* | 47:23 (67.14%) | 20:16 (55.56%) | 14:16 (46.67%) | .137 |
| *Education* | 15.61 (2.88) | 17.03 (2.88) | 16.91 (3.30) | .031* |
| **Cognition** |  |  |  |  |
| *CDR box score* | 6.3 (3.3) | 4.1 (2.3) | 2.5 (2.5) | <.001* |
| *CDR language* | 0.7 (0.6) | 1.1 (0.6) | 1.3 (0.7) | <.001* |
| *CDR behavior* | 1.3 (0.8) | 1.1 (0.6) | 0.5 (0.5) | <.001* |
| *MMSE* | 23.6 (4.9) | 24.4 (5.1) | 25.5 (5.1) | .287 |
| *CVLT memory* | 19.3 (7.7) | 17.0 (6.5) | 21.9 (7.7) | .053 |
| *CVLT recall after 30second delay* | 4.3 (2.7) | 2.9 (2.5) | 5.5 (2.9) | .002* |
| *CVLT recall after 10minute delay* | 3.2 (2.8) | 1.8 (2.3) | 5.3 (2.8) | <.001* |
| *CVLT recognition* | 7.1 (1.8) | 6.4 (2.4) | 8.2 (0.9) | .002* |
| *Digit span forward* | 5.6 (1.3) | 6.8 (1.5) | 5.0 (1.3) | <.001* |
| *Digit span backward* | 3.5 (1.4) | 4.9 (1.3) | 3.5 (1.4) | <.001* |
| *MTMT correct lines* | 9.9 (5.0) | 13.3 (2.5) | 12.2 (3.9) | .001* |
| *MTMT time* | 74.0 (40.4) | 46.9 (29.5) | 62.9 (39.1) | .008* |
| *Verbal fluency phonological* | 6.6 (4.5) | 8.6 (4.5) | 7.0 (5.0) | .132 |
| *Verbal fluency semantic* | 9.4 (6.3) | 8.1 (4.1) | 11.6 (8.4) | .137 |
| *BNT* | 12.2 (3.1) | 5.0 (3.6) | 12.3 (2.9) | <.001* |
| *PPVT* | 13.2 (3.3) | 8.6 (4.2) | 14.5 (2.0) | <.001* |

**Table 2.**
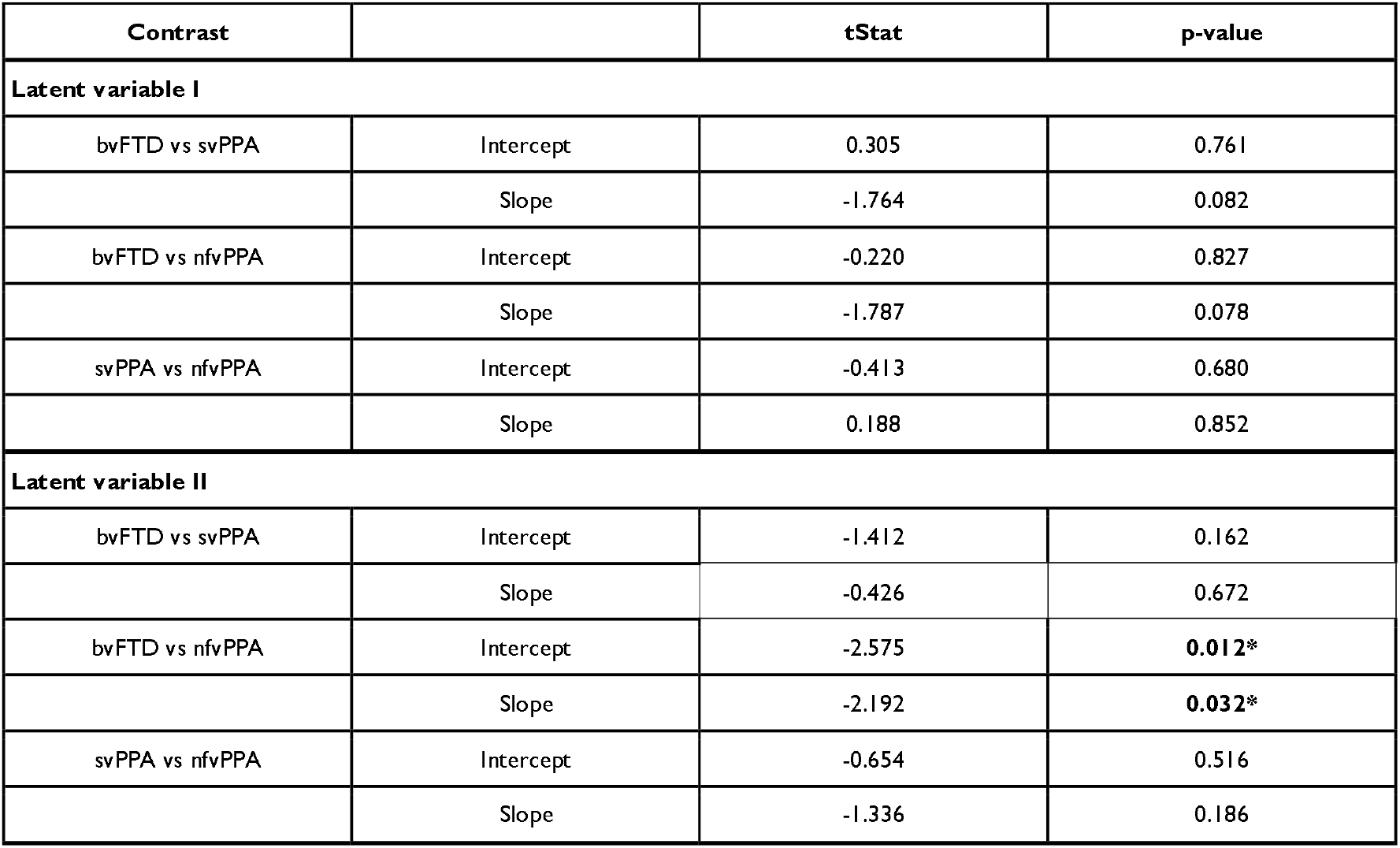
Differences in the predictions of the PLS model for the different FTD subtypes in LVs I and II, as determined by a linear regression model. P-values are reported after correction for multiple comparisons using a false discovery rate controlling method with a significance threshold of 0.05. Significant intercept or slope differences are indicated by an asterisk. bvFTD = behavioral variant FTD; svPPA = semantic variant PPA; nfvPPA = nonfluent variant PPA.

| Contrast |  | tStat | p-value |
| --- | --- | --- | --- |
| **Latent variable I** |  |  |  |
| bvFTD vs svPPA | Intercept | 0.305 | 0.761 |
|  | Slope | -1.764 | 0.082 |
| bvFTD vs nfvPPA | Intercept | -0.220 | 0.827 |
|  | Slope | -1.787 | 0.078 |
| svPPA vs nfvPPA | Intercept | -0.413 | 0.680 |
|  | Slope | 0.188 | 0.852 |
| **Latent variable II** |  |  |  |
| bvFTD vs svPPA | Intercept | -1.412 | 0.162 |
|  | Slope | -0.426 | 0.672 |
| bvFTD vs nfvPPA | Intercept | -2.575 | **0.012\*** |
|  | Slope | -2.192 | **0.032\*** |
| svPPA vs nfvPPA | Intercept | -0.654 | 0.516 |
|  | Slope | -1.336 | 0.186 |

### PLS analysis

The PLS analysis revealed four statistically significant latent variables relating behavioral measures in FTD to their corresponding brain atrophy patterns (permuted *p* < 0.05) at baseline. These patterns respectively account for 44.16%, 28.05%, 8.02%, and 5.75% (total of 85.98%) of the shared covariance between clinical and brain atrophy measures (Fig.2).

**Figure 2:**
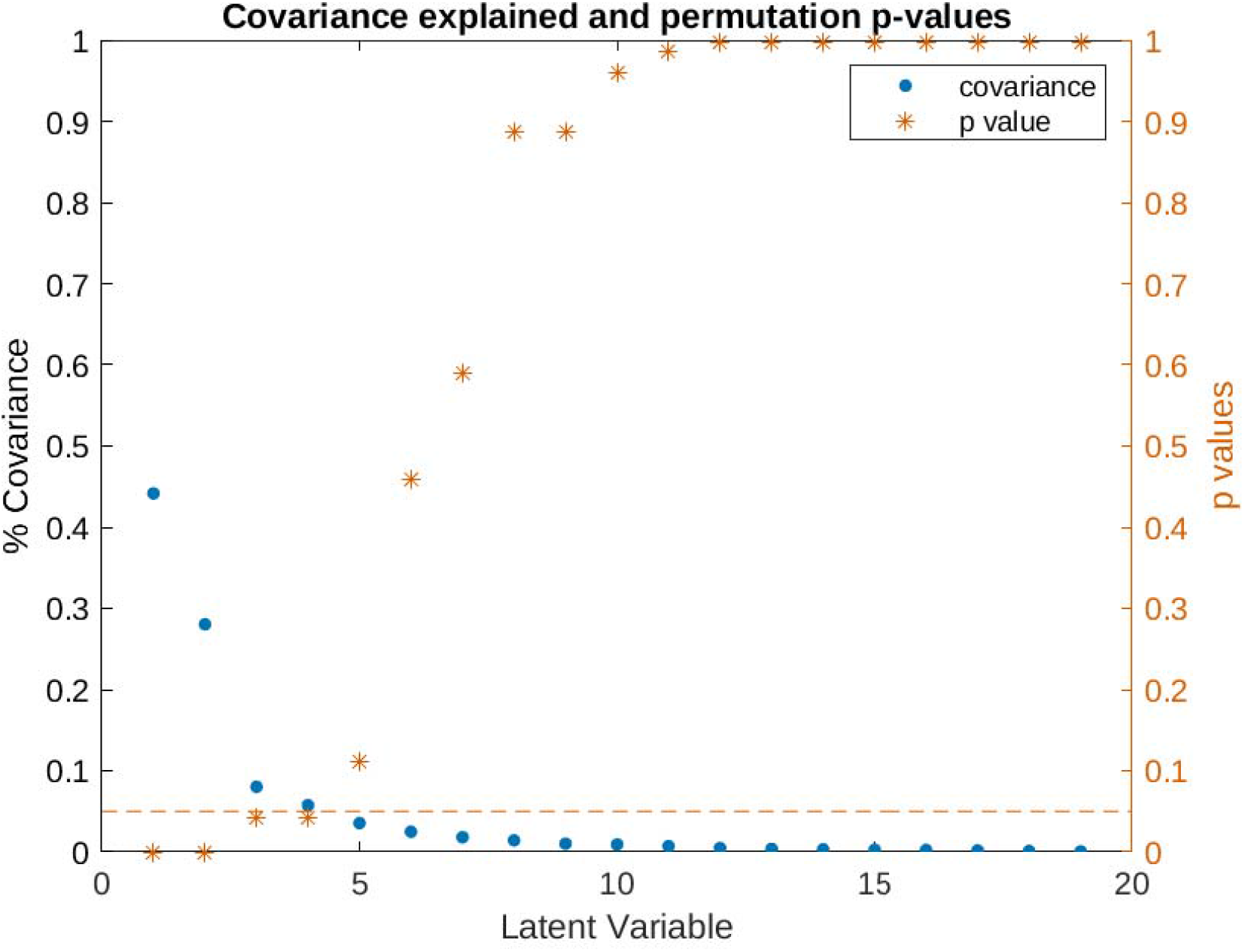
Covariance explained and permutation p-values for all latent variables in the PLS analysis. Latent variable I (LV-I, covariance explained = 44.16%, *p*<0.001), latent variable II (LV-II, covariance explained = 28.05%, *p*<0.001), latent variable III (LV-III, covariance explained = 8.02%, *p*<0.05), and latent variable IV (LV-IV, covariance explained = 5.75%, *p*<0.05) are selected for further analysis based on p-value (permuted *p* < 0.05).

### Behavioral and atrophy patterns

In summary, LV-I mostly represented confrontational naming and verbal learning/memory skills and was associated with temporal lobe structure and subcortical areas. LV-II captured diverse cognitive features in combination with frontal and subcortical atrophy patterns, while LV-III and LV-IV encompassed behavioral aspects and distributed atrophy networks.

The cognitive features contributing to the first LV (LV-I, Fig.3C), in order of magnitude, were impaired performance in confrontational naming (BNT and PPVT), verbal learning (CVLT), semantic verbal fluency, and MMSE, but comparatively higher scores in working memory/executive function (digit span). The corresponding brain pattern for this clinical profile was largely driven by atrophy in the temporal lobes and subcortical areas (amygdala, nucleus accumbens, hippocampus) on the one hand, and higher volume in the cerebellum on the other hand (Fig.3A/B, see *Supplementary Table* 2 and 3 for specific brain areas).

**Figure 3:**
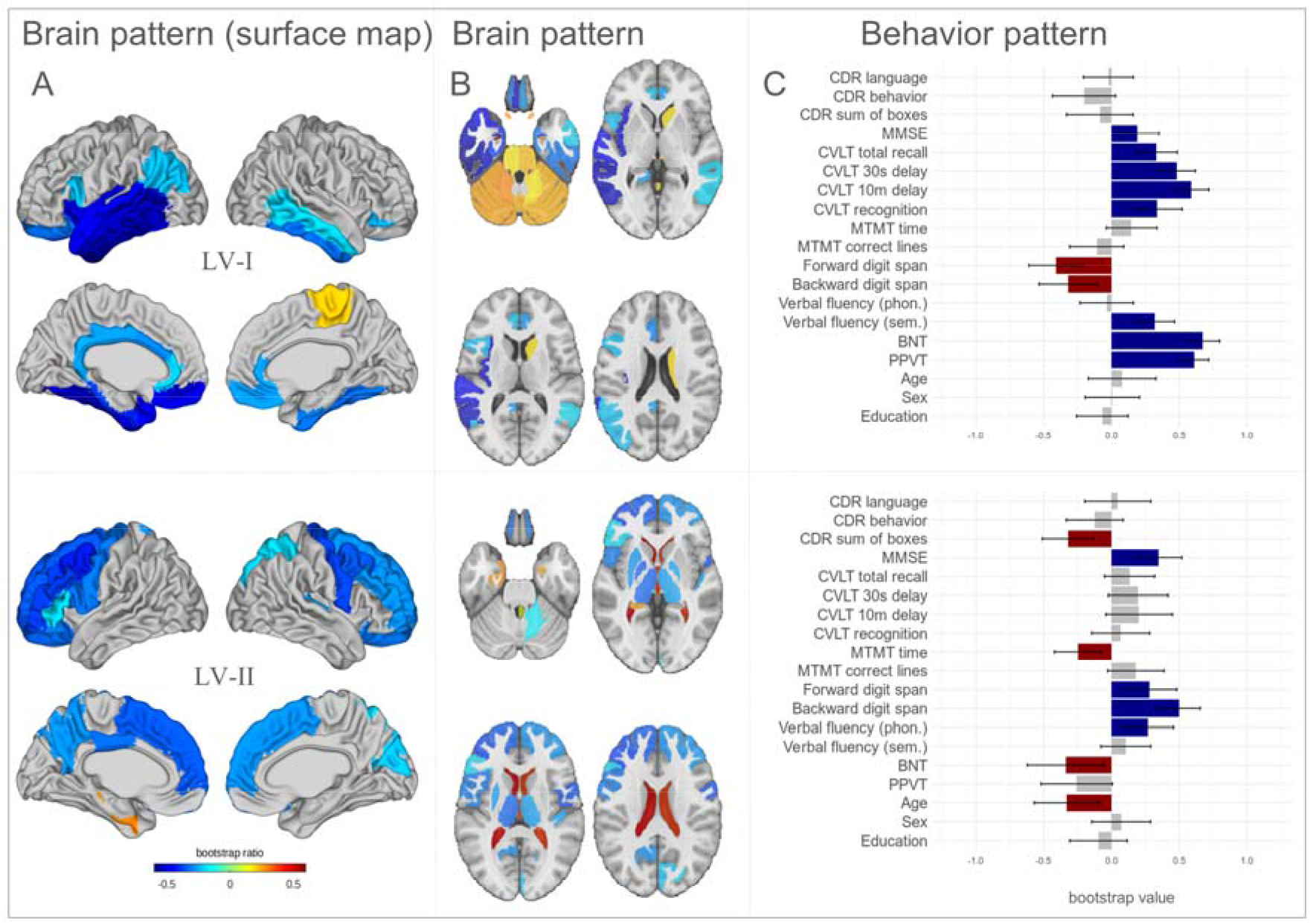
Latent variables I and II (LVs I - II) obtained from the PLS analysis. A, Brain pattern bootstrap ratios in MNI space, surface projection. Maps only include regions that significantly contribute to the LV, as estimated by bootstrapping values and confidence intervals. B, Brain pattern bootstrap values in MNI space, horizontal view illustrating subcortical structures and cerebellum. C, Pattern of demographic and cognitive test scores. The effect size estimates are derived from SVD analysis and the Confidence Intervals (CI) are calculated by bootstrapping.

For the second LV (LV-II), greater impairment in CDR sum of boxes, higher age, and higher BNT score, as well as lower MMSE scores, digit span, and phonological verbal fluency contributed to covariation. Regarding DBM measurements, predominantly increased ventricular expansion and lower volume in left frontal areas (including Broca’s area) and subcortical structures (pallidum, thalamus, but lower hippocampal volume), were involved.

*Supplementary Figure* 1 shows an example of how the putative brain network and the associated behavioral phenotype relate to each other. For each LV, we estimated patient-specific scores by projecting the brain and behavior patterns onto individual patients’ data (see Methods). The resulting scalar values (termed brain scores and behavior scores) reflect the extent to which an individual patient expresses each pattern. The two scores are correlated, i.e. patients with greater atrophy in the network in Fig.3A/B also tend to conform more closely to the clinical phenotype in Fig.3C. In LVs I and II, patients who score highly on both likely have more severe pathology, and we illustrate this by coloring the points (individual patients) by their MMSE scores. Individuals with more pronounced atrophy and symptom severity also tend to score highly on MMSE, a measure of global cognition.

Brain and behavior patterns for LVs III and IV are shown and discussed in the *Supplementary Figure* 2 and *Table* 4.

### Group differences

Fig.4 shows how the putative atrophy network and associated behavioral patterns are related to each other in individual patients and how these associations differ between FTD variants. While this measure is derived from the overall population, when compared between the groups of interest it can provide further insight regarding the heterogeneity and nuances of the disease sub-groups as it indicates whether the relationship and the degree of pattern expression are consistent or diverse across groups.

**Figure 4:**
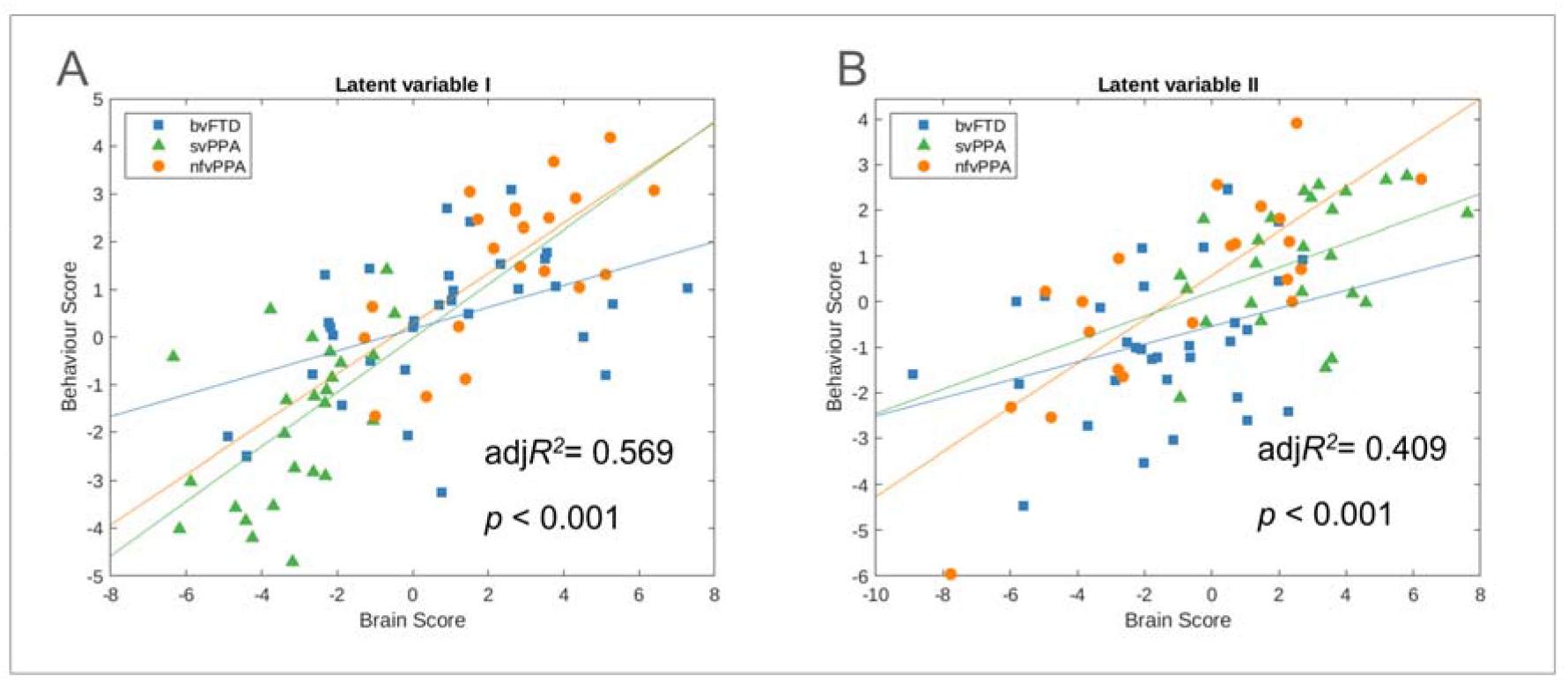
Individual patients’ brain versus behavioral PLS score and group differences between FTD variants. **A**, LV-I (adjusted *R*^2^= 0.569, *p*<.001). **B**, LV-II (adjusted *R*^2^ = 0.409, *p*<0.001). bvFTD = behavioral variant FTD; svPPA = semantic variant primary progressive aphasia; nfvPPA = nonfluent variant primary progressive aphasia.

Compared to bvFTD, the brain and behaviour scores in LV-I had stronger associations (i.e. steeper slopes) in svPPA and nfvPPA (Fig.4A), although these slope differences were statistically marginal (*p*∼0.08). For LV-II, the nfvPPA subtype had a significantly higher intercept (*tStat*=2.57, *p*=0.01) and slope (*tStat*=2.19, *p*=0.03) compared to the bvFTD variant, suggesting that it expresses the behaviour pattern to a greater extent and that the atrophy pattern has a more pronounced impact on cognition patterns in patients with nfvPPA, i.e. LV-II is reflective of the cognition and atrophy patterns of nfvPPA.

### Predictive capacity of the model

We examined the capacity of the combination of neural and behavioral patterns in the latent variables identified in the PLS analysis as features to predict clinical outcomes in patients. We applied a multi-class machine learning classifier with a 10-fold cross-validation loop on 100 randomized train and test splits. The predicted FTD subtype classification based on brain and behavior scores (including 16 cognitive scores and age, education, and sex) was then compared to clinician variant diagnoses; the resulting 3-class mean prediction accuracy over all repetitions was 89.12%. The mean sensitivity ranged from 84.19% for nfvPPA to 93.56% for bvFTD, with mean specificities of 91.46% for bvFTD to 97.15% for svPPA (Table 3). When we exclusively included brain scores in our model, the accuracy was reduced to 69.76%, whereas predictions based on the full battery of behavioral scores were 86.05% accurate.

**Table 3.** Results of the classification analysis. Machine learning was used to predict diagnosis of FTD variant for each participant in a k-fold cross-validation. Results are presented as mean values and SD. bvFTD = behavioral variant FTD; svPPA = semantic variant PPA; nfvPPA = nonfluent variant PPA.

|  | Classification accuracy | Sensitivity | Specificity | Balanced accuracy |
| --- | --- | --- | --- | --- |
| **Brain patterns only (including age/sex/education)** |  |  |  |  |
| brain | 69.76% (sd=2.73%) | bvFTD: 63.50% (sd=4.28%)<br>svPPA: 93.76% (sd=2.00%)<br>nfvPPA: 50.71% (sd=6.51%) | bvFTD: 76.28% (sd=3.15%)<br>svPPA: 94.92% (sd=1.65%)<br>nfvPPA: 82.47% (sd=2.26%) | bvFTD: 69.89%<br>svPPA: 94.34%<br>nfvPPA: 66.59% |
| in CDR matched sample brain | 68.99% (sd=2.78%) | bvFTD: 58.20% (sd=5.19%)<br>svPPA: 93.42% (sd=2.07%)<br>nfvPPA: 53.15% (sd=6.26%) | bvFTD: 77.39% (sd=2.97%)<br>svPPA: 93.40% (sd=1.91%)<br>nfvPPA: 82.69% (sd=2.18%) | bvFTD: 67.80%<br>svPPA: 93.41%<br>nfvPPA: 67.92% |
| longitudinal data brain | 71.94% (sd=3.08%) | bvFTD: 67.92% (sd=6.08%)<br>svPPA: 89.27% (sd=3.51%)<br>nfvPPA: 56.11% (sd=6.19%) | bvFTD: 79.35% (sd=3.46%)<br>svPPA: 94.95% (sd=1.14%)<br>nfvPPA: 83.52% (sd=3.05%) | bvFTD: 73.64%<br>svPPA: 92.11%<br>nfvPPA: 69.81% |
| **Behavior patterns only: Maximal model (including 16 cognitive scores, age/sex/education)** |  |  |  |  |
| behavior | 86.05% (sd=1.51%) | bvFTD: 87.34% (sd=2.49%)<br>svPPA: 84.92% (sd=2.12%)<br>nfvPPA: 85.43% (sd=2.61%) | bvFTD: 91.65% (sd=1.63%)<br>svPPA: 94.53% (sd=0.57%)<br>nfvPPA: 92.74% (sd=1.54%) | bvFTD: 89.50%<br>svPPA: 89.73%<br>nfvPPA: 89.08% |
| in CDR matched sample behavior | 85.88% (sd=1.41%) | bvFTD: 84.20% (sd=3.08%)<br>svPPA: 84.42% (sd=2.19%)<br>nfvPPA: 89.75% (sd=1.10%) | bvFTD: 93.66% (sd=1.30%)<br>svPPA: 93.40% (sd=0.38%)<br>nfvPPA: 91.88% (sd=1.64%) | bvFTD: 88.93%<br>svPPA: 88.91%<br>nfvPPA: 90.82% |
| longitudinal data behavior | 92.47% (sd=1.99%) | bvFTD: 92.00% (sd=1.64%)<br>svPPA: 86.82% (sd=5.69%)<br>nfvPPA: 100% (sd=0%) | bvFTD: 100% (sd=0%)<br>svPPA: 100% (sd=0%)<br>nfvPPA: 89.52% (sd=2.77%) | bvFTD: 96.00%<br>svPPA: 93.41%<br>nfvPPA: 94.76% |
| **Combination brain + behavior: Maximal model (including 16 cognitive scores, age/sex/education)** |  |  |  |  |
| brain + behavior | 89.12% (sd=1.21%) | bvFTD: 88.88% (sd=2.10%)<br>svPPA: 93.56% (sd=2.04%)<br>nfvPPA: 84.19% (sd=3.02%) | bvFTD: 91.46% (sd=1.75%)<br>svPPA: 97.15% (sd=1.06%)<br>nfvPPA: 94.65% (sd=0.80%) | bvFTD: 90.17%<br>svPPA: 95.36%<br>nfvPPA: 89.42% |
| in CDR matched sample brain + behavior | 88.88% (sd=1.51%) | bvFTD: 85.84% (sd=2.63%)<br>svPPA: 93.25% (sd=2.12%)<br>nfvPPA: 87.45% (sd=2.80%) | bvFTD: 92.89% (sd=1.64%)<br>svPPA: 96.78% (sd=1.16%)<br>nfvPPA: 93.69% (sd=0.77%) | bvFTD: 89.37%<br>svPPA: 95.02%<br>nfvPPA: 90.57% |
| longitudinal data brain + behavior | 88.09% (sd=1.45%) | bvFTD: 85.71% (sd=3.43%)<br>svPPA: 92.31% (sd=1.56%)<br>nfvPPA: 90.00% (sd=0%) | bvFTD: 95.65% (sd=0.86%)<br>svPPA: 95.83% (sd=0.48%)<br>nfvPPA: 92.59% (sd=1.82%) | bvFTD: 90.68%<br>svPPA: 94.07%<br>nfvPPA: 91.30% |
| **Behavior only: Minimal model (including CDR, BNT, age/sex/education)** |  |  |  |  |
| behavior (CDR + BNT) | 76.38% (sd=1.20%) | bvFTD: 84.82% (sd=1.80%)<br>svPPA: 70.19% (sd=1.74%)<br>nfvPPA: 61.13% (sd=3.08%) | bvFTD: 80.91% (sd=1.74%)<br>svPPA: 89.07% (sd=0.82%)<br>nfvPPA: 91.52% (sd=1.23%) | bvFTD: 82.87%<br>svPPA: 79.63%<br>nfvPPA: 76.33% |
| CDR | 63.87% (sd=1.70%) | bvFTD: 83.14% (sd=1.79%)<br>svPPA: 25.58% (sd=3.07%)<br>nfvPPA: 65.52% (sd=4.27%) | bvFTD: 64.48% (sd=1.53%)<br>svPPA: 83.52% (sd=1.93%)<br>nfvPPA: 91.26% (sd=0.81%) | bvFTD: 73.81%<br>svPPA: 54.55%<br>nfvPPA: 78.39% |
| BNT | 71.77% (sd=0.99%) | bvFTD: 84.75% (sd=1.30%)<br>svPPA: 78.12% (sd=0.00%)<br>nfvPPA: 28.13% (sd=3.47%) | bvFTD: 65.41% (sd=1.64%)<br>svPPA: 89.97% (sd=0.37%)<br>nfvPPA: 93.96% (sd=1.04%) | bvFTD: 75.08%<br>svPPA: 84.05%<br>nfvPPA: 61.05% |
| in CDR matched sample behavior | 73.98% (sd=1.42%) | bvFTD: 80.00% (sd=2.37%)<br>svPPA: 71.53% (sd=1.86%)<br>nfvPPA: 63.91% (sd=3.22%) | bvFTD: 81.73% (sd=1.84%)<br>svPPA: 87.86% (sd=1.02%)<br>nfvPPA: 89.67% (sd=1.48%) | bvFTD: 80.87%<br>svPPA: 79.70%<br>nfvPPA: 76.79% |
| in CDR matched sample CDR | 60.10% (sd=1.94%) | bvFTD: 78.23% (sd=2.06%)<br>svPPA: 27.09% (sd=3.25%)<br>nfvPPA: 66.74% (sd=4.56%) | bvFTD: 67.07% (sd=1.63%)<br>svPPA: 79.62% (sd=2.41%)<br>nfvPPA: 89.65% (sd=0.94%) | bvFTD: 72.65%<br>svPPA: 53.36%<br>nfvPPA: 78.20% |
| in CDR matched sample BNT | 70.17% (sd=0.99%) | bvFTD: 83.31% (sd=0.89%)<br>svPPA: 80.00% (sd=0.00%)<br>nfvPPA: 29.35% (sd=3.62%) | bvFTD: 65.34% (sd=1.73%)<br>svPPA: 88.99% (sd=0.45%)<br>nfvPPA: 94.77% (sd=0.86%) | bvFTD: 74.33%<br>svPPA: 84.50%<br>nfvPPA: 62.06% |
| longitudinal data<br>behavior | 84.95% (sd=1.22%) | bvFTD: 85.08% (sd=1.72%)<br>svPPA: 89.05% (sd=1.97%)<br>nfvPPA: 79.13% (sd=2.25%) | bvFTD: 91.40% (sd=1.18%)<br>svPPA: 90.36% (sd=0.91%)<br>nfvPPA: 95.03% (sd=1.28%) | bvFTD: 88.24%<br>svPPA: 89.71%<br>nfvPPA: 87.08% |
| **Combination brain + behavior: Minimal model (including CDR, BNT, age/sex/education)** |  |  |  |  |
| brain + behavior | 83.62% (sd=1.19%) | bvFTD: 84.35% (sd=1.61%)<br>svPPA: 87.50% (sd=0.00%)<br>nfvPPA: 76.17% (sd=4.21%) | bvFTD: 86.38% (sd=1.76%)<br>svPPA: 93.34% (sd=0.79%)<br>nfvPPA: 93.51% (sd=0.89%) | bvFTD: 85.37%<br>svPPA: 90.42%<br>nfvPPA: 84.84% |
| CDR + brain | 73.63% (sd=1.40%) | bvFTD: 80.46% (sd=1.53%)<br>svPPA: 64.78% (sd=2.73%)<br>nfvPPA: 68.38% (sd=3.80%) | bvFTD: 77.17% (sd=1.83%)<br>svPPA: 87.20% (sd=1.14%)<br>nfvPPA: 92.43% (sd=0.99%) | bvFTD: 78.82%<br>svPPA: 75.99%<br>nfvPPA: 80.40% |
| BNT + brain | 76.74% (sd=1.16%) | bvFTD: 83.72% (sd=1.37%)<br>svPPA: 87.84% (sd=1.32%)<br>nfvPPA: 43.04% (sd=3.40%) | bvFTD: 69.27% (sd=1.82%)<br>svPPA: 95.33% (sd=0.47%)<br>nfvPPA: 93.02% (sd=0.99%) | bvFTD: 76.50%<br>svPPA: 91.59%<br>nfvPPA: 68.03% |
| in CDR matched sample<br>brain + behavior | 81.54% (sd=1.42%) | bvFTD: 79.24% (sd=2.13%)<br>svPPA: 90.00% (sd=0.00%)<br>nfvPPA: 75.14% (sd=4.40%) | bvFTD: 87.54% (sd=1.86%)<br>svPPA: 91.75% (sd=0.98%)<br>nfvPPA: 92.03% (sd=1.09%) | bvFTD: 83.39%<br>svPPA: 90.88%<br>nfvPPA: 83.59% |
| in CDR matched sample<br>CDR + brain | 72.39% (sd=1.60%) | bvFTD: 76.38% (sd=1.73%)<br>svPPA: 68.35% (sd=2.61%)<br>nfvPPA: 69.78% (sd=4.10%) | bvFTD: 80.46% (sd=1.87%)<br>svPPA: 85.65% (sd=1.32%)<br>nfvPPA: 90.77% (sd=1.21%) | bvFTD: 78.42%<br>svPPA: 77.00%<br>nfvPPA: 80.28% |
| in CDR matched sample<br>BNT + brain | 75.08% (sd=1.22%) | bvFTD: 81.92% (sd=1.45%)<br>svPPA: 90.37% (sd=1.41%)<br>nfvPPA: 40.57% (sd=3.55%) | bvFTD: 69.42% (sd=1.92%)<br>svPPA: 94.24% (sd=0.57%)<br>nfvPPA: 93.59% (sd=1.01%) | bvFTD: 75.67%<br>svPPA: 92.31%<br>nfvPPA: 67.08% |
| longitudinal data<br>brain + behavior | 85.77% (sd=2.04%) | bvFTD: 81.77% (sd=2.64%)<br>svPPA: 92.85% (sd=2.59%)<br>nfvPPA: 87.00% (sd=5.71%) | bvFTD: 94.43% (sd=2.45%)<br>svPPA: 90.27% (sd=1.33%)<br>nfvPPA: 94.38% (sd=1.72%) | bvFTD: 88.10%<br>svPPA: 91.56%<br>nfvPPA: 90.69% |

Projecting the results of the PLS analysis onto the longitudinal data of the same cohort as an in-sample validation (with subject-level cross-validation), our classification model achieved an accuracy of 88.09%, suggesting robustness of our model.

To assess whether our model was adaptable for a clinical setting with limited time and resources, we repeated our analyses while only including the CDR scales and the BNT into the PLS and our classification model (*Supplementary Figure 3*). Using these minimal variables, our model still achieved an accurate FTD subtype classification of 83.62%. Here, the combination of brain and clinical measures was crucial as predictions only using the behavior scores were only accurate 76.38% of the time. Notably, adding MRI measures to the clinical model increased the sensitivity for both PPAs by over 15% (70.19% to 87.50% for svPPA and 61.13% to 76.17% for nfvPPA).

To ensure our models did not solely rely on disease severity as a predictor, we repeated all the analyses in severity-matched subsamples (CDR<1.5). The models achieved similar accuracies within these severity-matched samples (Table 3), suggesting that the models do not use severity in symptoms to differentiate the patients across variants.

### Stability of brain atrophy and clinical patterns

To ensure that age, sex, or education were not driving the observed relationship between brain characteristics and cognitive function in the patients, we repeated the analyses after regressing out the effects of age, sex, and education based on the healthy control group (see *Supplementary Table* 5). The results remained largely unchanged in terms of the number of LVs, their behavioral patterns and differences between FTD variants, and their prediction accuracy in classifying subjects into FTD subgroups. This suggests that the findings in FTD patients reflect disease processes rather than changes in cognitive functioning in healthy aging. Neural patterns were very similar after this step, as shown in *Supplementary Figure 4*.

### Longitudinal progression of brain atrophy and clinical patterns

In terms of the longitudinal changes of brain and behavior scores in the entire patient cohort, we found that the brain patterns in all four LVs (I: *p*=0.007, II: *p*<0.001, III: *p*<0.001, IV: *p*<0.003) as well as behavior patterns in LVs I and II (I: *p*=0.006, II: *p*<0.002) significantly worsened over time (*Supplementary Figure 5*). Scores in the behavioral pattern of LV-IV slightly increased over time (*p*=0.007). As expected, since baseline and follow-up visits were only about one year apart, the changes are relatively small. With regard to the yearly rate of change in the three diagnostic groups, we only found differences between variants in LV-I (Fig.5). Here, we found significant differences in the delta values for behavioral scores, with increased changes in svPPA individuals compared to both bvFTD (*tStat*=-2.8541, *p*<0.01) and nfvPPA (*tStat*=-3.1358, *p*<0.006). Similarly, the svPPA variant changed more in terms of their brain scores than the nfvPPA subtype (*tStat*=-3.2975, *p*=0.004).

**Figure 5:**
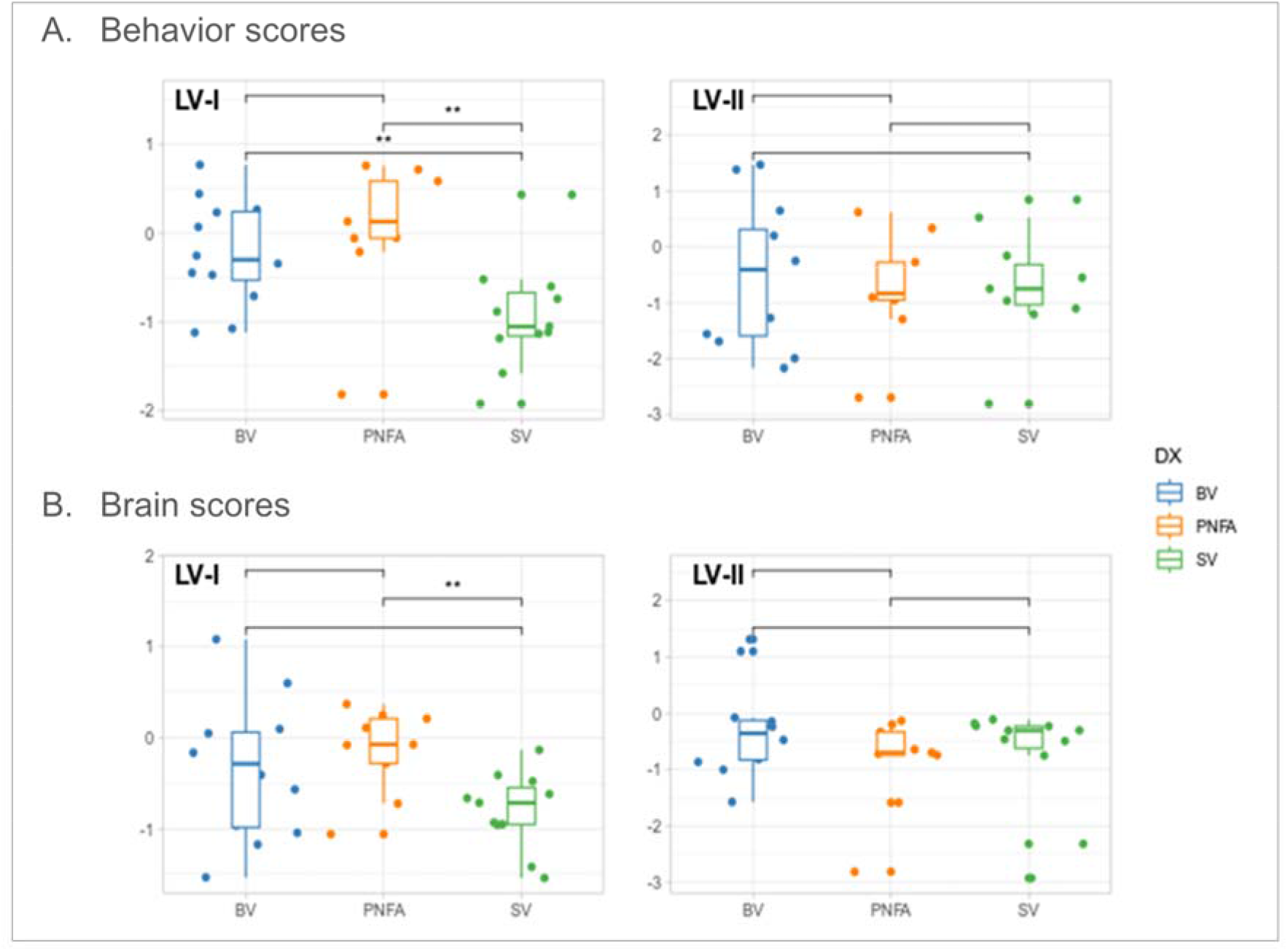
Boxplots showing the longitudinal progression of brain and behavior scores for LVs I and II. A, Yearly rate of change in behavior scores in the three FTD variants. B, Yearly rate of change in brain scores in the three FTD variants. Asterisks indicate significant group differences based on unpaired t-tests comparing the groups. BV: bvFTD, SV: svPPA, PNFA: nfvPPA.

## Discussion

The present study links clinical features of the three core FTD subtypes, bvFTD, svPPA, and nfvPPA, to the underlying brain atrophy pattern using a single integrated analysis. In these three cohorts, in addition to higher age, a range of cognitive characteristics, including global cognitive function, language, and executive function, were linked to brain atrophy. While atrophy patterns were widely distributed across both cortical and subcortical structures, the covariance between atrophy and behavioral measures was largely explained by the involvement of frontal and temporal lobes and subcortical structures. We also found differences in baseline and longitudinal cognitive performance and in the relationship between atrophy and cognition between bvFTD, svPPA, and nfvPPA. This allowed us to use the observed atrophy and cognition patterns in a prediction model, yielding high prediction accuracy (89.12%), sensitivity, and specificity in classifying FTD patients into phenotypes as compared to clinician subtype diagnosis as a gold standard. Even when only including minimal features (DBM values, CDR subscales, and BNT scores) as predictors, our model outperformed previous attempts at automated diagnosis of FTD, thereby demonstrating clinical utility. In this scenario, the MRI-derived features crucially contributed to the diagnostic accuracy (76% versus 83% without and with MRI features, respectively). This provides promising evidence that the combination of DBM and multivariate statistical methods could potentially aid the automated diagnosis and classification of FTD patients, in addition to constituting a means to investigate the complex manifestations of cognitive decline in FTD in relation to patterns of brain atrophy.

Our data driven PLS results align with previous findings on neuropathological changes in FTD. We found that the covariation of FTD groups explained in LV-I was partially explained by atrophy in the temporal lobe. While temporal lobe atrophy has been mostly associated with svPPA^12^, some studies indicate that it can also contribute to symptoms in bvFTD, especially in combination with hippocampal abnormalities^56^, which we also saw in LV-II. This has been interpreted as the involvement of dysfunction of the default mode, limbic, and salience networks in the pathogenesis of bvFTD^57^, leading to the observed changes in emotional and social processing. Particularly in LV-II, our results show that atrophy in the frontal lobe plays a crucial role in behavioral and cognitive symptoms in FTD, including impairment of executive function, attention, and language. Additionally, we found that structural changes in the insula contributed to cognitive decline and specifically seemed todistinguish nfvPPA from svPPA in LV-I. Due to its extensive anatomical and functional connections to linguistic, motor, and sensory areas in the frontal cortex^58,59^, the insula has been associated with semantic, syntactic, and phonological processing^60^ and seems to be responsible for the development of motor speech disorders like apraxia^61^. This explains its potential involvement in the pathophysiology of nfvPPA. The atrophy pattern in LV-III was largely driven by cerebellar structural changes. Behavioral impairment in genetic FTD has been associated with cerebellar atrophy^56,62^ and seems to show distinct patterns between FTD subtypes, specifically involvement of lobule VII in bvFTD and lobule VI in nfvPPA^63^ which we found in LV-I, although we did not find focal atrophy in lobule V as previously described^63^. While the contribution of frontotemporal-lobar degeneration to cognitive symptoms in FTD has been widely established^5^, subcortical atrophy is only recently receiving more attention. In LV-II, we found pronounced atrophy around the ventricles as distinctive features between FTD subgroups, confirming findings that ventricular expansion is a common feature of bvFTD and constitutes a sensitive and reliable marker of disease progression^29^. Involvement of the thalamus and amygdala, which we found in several LVs, has been reported for both sporadic and genetic FTD and seems to precede cortical atrophy^56,57,64^. Previous studies have shown distinctive atrophy patterns in the thalamus between the core FTD subtypes^65,66^. Considering that the amygdala is part of the limbic system and is implicated in emotional processing and reward learning^67^, it is not surprising that our analyses found involvement of this structure. Overall, this suggests that investigating subcortical atrophy in FTD is a promising avenue for both diagnostic purposes and improving our understanding of neural mechanisms underlying clinical presentation. It also highlights the utility of DBM in assessing cortical and subcortical atrophy given its improved accuracy in detecting structural changes in deeper brain layers.

To test whether the brain and behaviour patterns derived from the PLS analysis were useful as informative features that could separate individual patients with different FTD syndromes, we assessed prediction of FTD subtype in an automated machine learning procedure. Classification accuracy was 89.12%, with sensitivities for different variants ranging from 84.19% to 93.56% and specificities of 91.46% to 97.15%. We validated the robustness of our model by projecting the results of the PLS analysis onto the longitudinal data of the same cohort as well as testing our model in a disease severity-matched sample of this cohort and obtained similar accuracies. Importantly, we also tested whether the combination of atrophy scores and only two neuropsychological tests (CDR and BNT) would be sufficient to distinguish FTD syndromes. These measures were chosen because they are clinically useful, and because they require little time compared to the full test battery used in our maximal model. This is relevant as patients’ attention and capability to endure lengthy examinations is reduced and clinicians have limited time for extensive testing. Even with these minimal input variables (combined with the MRI information), our model achieved an accuracy of 83.62%.

These accuracies are comparable with or superior to those of similar studies. While studies usually achieve high accuracy when differentiating FTD patients from healthy controls^10^, few groups attempted to identify the FTD subtype^22,41^, only one of which used a three-way comparison rather than comparing two groups against each other at a time^42^. The latter study resulted in high specificity (94.2-97.5%) but low sensitivity (0-50%). Given that predictions were less accurate in our study when we used behavioral variables only, the strength of our approach seems to lie in the combination of neural and clinical characteristics in the patients, particularly the use of DBM, as well as the application of a multivariate statistics method that allows us to capture the maximal covariance across features that leads to a data-driven separation between groups, indicated by the clear separation between variants in terms of brain scores in LVs I and IV particularly. These results suggest that automated methods incorporating DBM-derived atrophy and clinical performance could assist in the diagnosis of FTD subtypes.

One of the strengths of this study is that our image processing pipelines have been developed and extensively validated for use in multi-center and multi-scanner datasets of aging and neurodegenerative disease populations and provide robust and sensitive DBM measures^29,30,53^. We further quality-controlled all the steps of the pipeline to ensure the accuracy of the results. Similarly, we have demonstrated the reliability of our analysis results repeating all steps after regressing out the effects of healthy aging. An intriguing result of this study is the high prediction accuracy (over 89%) our approach achieved for classifying FTD patients according to FTD variant. This is particularly relevant given that differential diagnosis of FTD as opposed to other psychiatric disorders and even controls still poses a challenge^9,68,69^. While many MRI studies indicate good diagnostic accuracy when comparing subjects with bvFTD and cognitively unimpaired individuals or patients with Alzheimer’s Disease, they rarely achieve a higher accuracy than about 80%^10^. To our knowledge, there is only one study to date that has attempted to predict FTD subtypes solely based on MRI data^70^. This study focused on ventricular features to discriminate bvFTD from PPAs in the FTLDNI cohort. The best accuracies in these dichotomous comparisons were 66% and 71% respectively, meaning our approach to a three-way comparison outperformed this study. This highlights the increased sensitivity of DBM as well as the potential utility of DBM paired with multivariate statistics in the diagnosis of FTD. This is an important step for improving patient care and diagnostic prognosis.

The present study has some limitations that need to be taken into account. Although PLS provides a comprehensive approach to investigating brain-behavior relations, it cannot provide insight into how each particular clinical manifestation potentially relates to a specific brain region, rather than the atrophy pattern as a whole. Such individual relationships need to be addressed in future studies. Another methodological consideration is that these results are valid only for the sample of FTD patients included here. Further validation is needed in larger, more diverse samples before we can be confident that the observed results will generalize to the rest of the population. These samples should include logopenic and semantic behavioral as well as genetic variants of FTD to cover the entire disease spectrum as well as subjects with a higher range of educational and ethnic backgrounds. Furthermore, sample sizes in the FTLDNI cohort are small, particularly for the two PPA groups, and some relevant information on the subjects is missing, such as disease duration, time from diagnosis, and comorbidities, potentially obscuring confounding factors in our analyses. It should also be noted that the bvFTD patients displayed higher disease severity than other groups which might impact our analyses. Similarly, our analyses could not cover all cognitive domains since many participants did not undergo respective testing and the FTLDNI did not collect information on language comprehension and writing/reading skills, for instance. To investigate the diagnostic utility of our approach which uses machine learning and multivariate statistics, future studies could directly compare traditional approaches, including cortical thickness and voxel-based morphometry, to our results and include genetic variants of FTD in their sample to ensure generalizability over the entire spectrum of FTD.

Altogether, findings in this study demonstrate a robust mapping between neurodegeneration as estimated by DBM values and the cognitive manifestations of the core FTD subtypes. The combination of DBM and multivariate statistical methods could potentially serve as an imaging biomarker for diagnosis and phenotyping in FTD and thereby improve early disease management and automated diagnosis.

## Data Availability

NIFD MRI and clinical measures are available through https://ida.loni.usc.edu/login.jsp. Derived data supporting the findings of this study, including individual PLS scores, are available from the corresponding author on request.

## Acknowledgments

The authors would like to thank Dr. Boris Bernhard, Dr. Bratislav Misic, Aliza Brzezinski-Rittner, Roqaie Moqaddam, Walter Adame-Gonzalez, Zaki Alasmar, Dr. Isabelle Lajoie, and Dr. Farooq Kamal for helpful discussions and feedback during this study. The authors also acknowledge Compute Canada (https://www.computecanada.ca/home) for the usage of the computing resources in the current work.

## References

1. Mackenzie IRA, Neumann M, Bigio EH, et al. Nomenclature for neuropathologic subtypes of frontotemporal lobar degeneration: consensus recommendations. Acta Neuropathol (Berl). 2009;117(1):15–18. doi:10.1007/s00401-008-0460-5

2. Mackenzie IRA, Neumann M, Bigio EH, et al. Nomenclature and nosology for neuropathologic subtypes of frontotemporal lobar degeneration: an update. Acta Neuropathol (Berl). 2010;119(1):1–4. doi:10.1007/s00401-009-0612-2

3. Rademakers R, Neumann M, Mackenzie IR. Advances in understanding the molecular basis of frontotemporal dementia. Nat Rev Neurol. 2012;8(8):423–434. doi:10.1038/nrneurol.2012.117

4. Bang J, Spina S, Miller BL. Frontotemporal dementia. The Lancet. 2015;386(10004):1672–1682. doi:10.1016/S0140-6736(15)00461-4

5. Olney NT, Spina S, Miller BL. Frontotemporal Dementia. Neurol Clin. 2017;35(2):339-374. doi:10.1016/j.ncl.2017.01.008

6. Murley AG, Coyle-Gilchrist I, Rouse MA, et al. Redefining the multidimensional clinical phenotypes of frontotemporal lobar degeneration syndromes. Brain. 2020;143(5):1555–1571. doi:10.1093/brain/awaa097

7. Rascovsky K, Hodges JR, Knopman D, et al. Sensitivity of revised diagnostic criteria for the behavioural variant of frontotemporal dementia. Brain. 2011;134(9):2456–2477. doi:10.1093/brain/awr179

8. Gorno-Tempini ML, Hillis AE, Weintraub S, et al. Classification of primary progressive aphasia and its variants. Neurology. 2011;76(11):1006–1014. doi:10.1212/WNL.0b013e31821103e6

9. Ducharme S, Price BH, Larvie M, Dougherty DD, Dickerson BC. Clinical Approach to the Differential Diagnosis Between Behavioral Variant Frontotemporal Dementia and Primary Psychiatric Disorders. Am J Psychiatry. 2015;172(9):827–837. doi:10.1176/appi.ajp.2015.14101248

10. McCarthy J, Collins DL, Ducharme S. Morphometric MRI as a diagnostic biomarker of frontotemporal dementia: A systematic review to determine clinical applicability. NeuroImage Clin. 2018;20:685–696. doi:10.1016/j.nicl.2018.08.028

11. Vijverberg EGB, Wattjes MP, Dols A, et al. Diagnostic Accuracy of MRI and Additional [18F]FDG-PET for Behavioral Variant Frontotemporal Dementia in Patients with Late Onset Behavioral Changes. J Alzheimers Dis. 2016;53(4):1287–1297. doi:10.3233/JAD-160285

12. Meeter LH, Kaat LD, Rohrer JD, Van Swieten JC. Imaging and fluid biomarkers in frontotemporal dementia. Nat Rev Neurol. 2017;13(7):406–419. doi:10.1038/nrneurol.2017.75

13. Chu M, Jiang D, Li D, et al. Atrophy network mapping of clinical subtypes and main symptoms in frontotemporal dementia. Brain. Published online March 1, 2024:awae067. doi:10.1093/brain/awae067

14. Pan PL, Song W, Yang J, et al. Gray Matter Atrophy in Behavioral Variant Frontotemporal Dementia: A Meta-Analysis of Voxel-Based Morphometry Studies. Dement Geriatr Cogn Disord. 2012;33(2-3):141–148. doi:10.1159/000338176

15. Rosen HJ, Allison SC, Schauer GF, Gorno-Tempini ML, Weiner MW, Miller BL. Neuroanatomical correlates of behavioural disorders in dementia. Brain. 2005;128(11):2612–2625. doi:10.1093/brain/awh628

16. Schroeter ML, Laird AR, Chwiesko C, et al. Conceptualizing neuropsychiatric diseases with multimodal data-driven meta-analyses – The case of behavioral variant frontotemporal dementia. Cortex. 2014;57:22–37. doi:10.1016/j.cortex.2014.02.022

17. Seeley WW, Crawford R, Rascovsky K, et al. Frontal Paralimbic Network Atrophy in Very Mild Behavioral Variant Frontotemporal Dementia. Arch Neurol. 2008;65(2). doi:10.1001/archneurol.2007.38

18. Chan D, Fox NC, Scahill RI, et al. Patterns of temporal lobe atrophy in semantic dementia and Alzheimer’s disease. Ann Neurol. 2001;49(4):433–442.

19. Davies RR, Graham KS, Xuereb JH, Williams GB, Hodges JR. The human perirhinal cortex and semantic memory. Eur J Neurosci. 2004;20(9):2441–2446. doi:10.1111/j.1460-9568.2004.03710.x

20. Davies RR, Halliday GM, Xuereb JH, Kril JJ, Hodges JR. The neural basis of semantic memory: Evidence from semantic dementia. Neurobiol Aging. 2009;30(12):2043–2052. doi:10.1016/j.neurobiolaging.2008.02.005

21. Galton CJ, Patterson K, Graham K, et al. Differing patterns of temporal atrophy in Alzheimer’s disease and semantic dementia. Neurology. 2001;57(2):216–225. doi:10.1212/WNL.57.2.216

22. Bisenius S, Neumann J, Schroeter ML. Validating new diagnostic imaging criteria for primary progressive aphasia via anatomical likelihood estimation meta□analyses. Eur J Neurol. 2016;23(4):704–712. doi:10.1111/ene.12902

23. Gorno□Tempini ML, Dronkers NF, Rankin KP, et al. Cognition and anatomy in three variants of primary progressive aphasia. Ann Neurol. 2004;55(3):335–346. doi:10.1002/ana.10825

24. Mesulam M, Wieneke C, Rogalski E, Cobia D, Thompson C, Weintraub S. Quantitative Template for Subtyping Primary Progressive Aphasia. Arch Neurol. 2009;66(12). doi:10.1001/archneurol.2009.288

25. Rogalski E, Cobia D, Martersteck A, et al. Asymmetry of cortical decline in subtypes of primary progressive aphasia. Neurology. 2014;83(13):1184–1191. doi:10.1212/WNL.0000000000000824

26. Dadar M, Potvin O, Camicioli R, Duchesne S, for the Alzheimer’s Disease Neuroimaging Initiative. Beware of white matter hyperintensities causing systematic errors in FreeSurfer gray matter segmentations! Hum Brain Mapp. 2021;42(9):2734–2745. doi:10.1002/hbm.25398

27. Ashburner J, Friston KJ. Voxel-Based Morphometry—The Methods. NeuroImage. 2000;11(6):805-821. doi:10.1006/nimg.2000.0582

28. Chung MK, Worsley KJ, Paus T, et al. A Unified Statistical Approach to Deformation-Based Morphometry. NeuroImage. 2001;14(3):595–606. doi:10.1006/nimg.2001.0862

29. Manera AL, Dadar M, Collins DL, Ducharme S. Deformation based morphometry study of longitudinal MRI changes in behavioral variant frontotemporal dementia. NeuroImage Clin. 2019;24:102079. doi:10.1016/j.nicl.2019.102079

30. Shafiei G, Bazinet V, Dadar M, et al. Network structure and transcriptomic vulnerability shape atrophy in frontotemporal dementia. Brain. 2023;146(1):321–336. doi:10.1093/brain/awac069

31. Wisse LEM, Ungrady MB, Ittyerah R, et al. Cross-sectional and longitudinal medial temporal lobe subregional atrophy patterns in semantic variant primary progressive aphasia. Neurobiol Aging. 2021;98:231–241. doi:10.1016/j.neurobiolaging.2020.11.012

32. Cardenas VA, Boxer AL, Chao LL, et al. Deformation-Based Morphometry Reveals Brain Atrophy in Frontotemporal Dementia. Arch Neurol. 2007;64(6):873. doi:10.1001/archneur.64.6.873

33. Scanlon C, Mueller SG, Tosun D, et al. Impact of Methodologic Choice for Automatic Detection of Different Aspects of Brain Atrophy by Using Temporal Lobe Epilepsy as a Model. Am J Neuroradiol. 2011;32(9):1669–1676. doi:10.3174/ajnr.A2578

34. Bron EE, Smits M, Papma JM, et al. Multiparametric computer-aided differential diagnosis of Alzheimer’s disease and frontotemporal dementia using structural and advanced MRI. Eur Radiol. 2017;27(8):3372–3382. doi:10.1007/s00330-016-4691-x

35. Dukart J, Mueller K, Horstmann A, et al. Combined Evaluation of FDG-PET and MRI Improves Detection and Differentiation of Dementia. Zhan W, ed. PLoS ONE. 2011;6(3):e18111. doi:10.1371/journal.pone.0018111

36. Kuceyeski A, Zhang Y, Raj A. Linking white matter integrity loss to associated cortical regions using structural connectivity information in Alzheimer’s disease and fronto-temporal dementia: The Loss in Connectivity (LoCo) score. NeuroImage. 2012;61(4):1311–1323. doi:10.1016/j.neuroimage.2012.03.039

37. Raamana PR, Rosen H, Miller B, Weiner MW, Wang L, Beg MF. Three-Class Differential Diagnosis among Alzheimer Disease, Frontotemporal Dementia, and Controls. Front Neurol. 2014;5. doi:10.3389/fneur.2014.00071

38. Wang J, Redmond SJ, Bertoux M, Hodges JR, Hornberger M. A Comparison of Magnetic Resonance Imaging and Neuropsychological Examination in the Diagnostic Distinction of Alzheimer’s Disease and Behavioral Variant Frontotemporal Dementia. Front Aging Neurosci. 2016;8. doi:10.3389/fnagi.2016.00119

39. Möller C, Hafkemeijer A, Pijnenburg YAL, et al. Joint assessment of white matter integrity, cortical and subcortical atrophy to distinguish AD from behavioral variant FTD: A two-center study. NeuroImage Clin. 2015;9:418–429. doi:10.1016/j.nicl.2015.08.022

40. Perovnik M, Vo A, Nguyen N, et al. Automated differential diagnosis of dementia syndromes using FDG PET and machine learning. Front Aging Neurosci. 2022;14:1005731. doi:10.3389/fnagi.2022.1005731

41. Wilson SM, Ogar JM, Laluz V, et al. Automated MRI-based classification of primary progressive aphasia variants. NeuroImage. 2009;47(4):1558–1567. doi:10.1016/j.neuroimage.2009.05.085

42. Tahmasian M, Shao J, Meng C, et al. Based on the Network Degeneration Hypothesis: Separating Individual Patients with Different Neurodegenerative Syndromes in a Preliminary Hybrid PET/MR Study. J Nucl Med. 2016;57(3):410–415. doi:10.2967/jnumed.115.165464

43. Kim JP, Kim J, Park YH, et al. Machine learning based hierarchical classification of frontotemporal dementia and Alzheimer’s disease. NeuroImage Clin. 2019;23:101811. doi:10.1016/j.nicl.2019.101811

44. McIntosh AR, Lobaugh NJ. Partial least squares analysis of neuroimaging data: applications and advances. NeuroImage. 2004;23 Suppl 1:S250–263. doi:10.1016/j.neuroimage.2004.07.020

45. McIntosh AR, Mišić B. Multivariate statistical analyses for neuroimaging data. Annu Rev Psychol. 2013;64:499–525. doi:10.1146/annurev-psych-113011-143804

46. Wold HOA. Nonlinear estimation by iterative least squares proce-dures. In: David, ed. Research Papers in Statistics, Festschrift for J.Neyman. Wiley; 1966:411-444.

47. Coupe P, Yger P, Prima S, Hellier P, Kervrann C, Barillot C. An Optimized Blockwise Nonlocal Means Denoising Filter for 3-D Magnetic Resonance Images. IEEE Trans Med Imaging. 2008;27(4):425–441. doi:10.1109/TMI.2007.906087

48. Sled JG, Zijdenbos AP, Evans AC. A nonparametric method for automatic correction of intensity nonuniformity in MRI data. IEEE Trans Med Imaging. 1998;17(1):87–97. doi:10.1109/42.668698

49. Dadar M, Fonov VS, Collins DL. A comparison of publicly available linear MRI stereotaxic registration techniques. NeuroImage. 2018;174:191–200. doi:10.1016/j.neuroimage.2018.03.025

50. Avants B, Tustison N, Gee J, Song G. ANTS: Advanced Open-Source Normalization Tools for Neuroanatomy. Published online 2009.

51. Fonov V, Evans AC, Botteron K, Almli CR, McKinstry RC, Collins DL. Unbiased average age-appropriate atlases for pediatric studies. NeuroImage. 2011;54(1):313–327. doi:10.1016/j.neuroimage.2010.07.033

52. Manera AL, Dadar M, Fonov V, Collins DL. CerebrA, registration and manual label correction of Mindboggle-101 atlas for MNI-ICBM152 template. Sci Data. 2020;7(1):237. doi:10.1038/s41597-020-0557-9

53. Zeighami Y, Fereshtehnejad SM, Dadar M, et al. A clinical-anatomical signature of Parkinson’s disease identified with partial least squares and magnetic resonance imaging. NeuroImage. 2019;190:69–78. doi:10.1016/j.neuroimage.2017.12.050

54. Eckart C, Young G. The approximation of one matrix by another of lower rank. Psychometrika. 1936;1(3):211–218. doi:10.1007/BF02288367

55. Efron B, Tibshirani R. Bootstrap Methods for Standard Errors, Confidence Intervals, and Other Measures of Statistical Accuracy. Stat Sci. 1986;1(1). doi:10.1214/ss/1177013815

56. Bussy A, Levy JP, Best T, et al. Cerebellar and subcortical atrophy contribute to psychiatric symptoms in frontotemporal dementia. Hum Brain Mapp. 2023;44(7):2684–2700. doi:10.1002/hbm.26220

57. Gordon E, Bocchetta M, Nicholas JM, Cash DM, Rohrer JD. Subcortical atrophy imaging biomarker performance across the sporadic and genetic frontotemporal dementias. Alzheimers Dement. 2022;18(S1):e062842. doi:10.1002/alz.062842

58. Augustine J. Circuitry and functional aspects of the insular lobe in primates including humans. Brain Res Rev. 1996;22(3):229–244. doi:10.1016/S0165-0173(96)00011-2

59. Jakab A, Molnár PP, Bogner P, Béres M, Berényi EL. Connectivity-based parcellation reveals interhemispheric differences in the insula. Brain Topogr. 2012;25(3):264–271. doi:10.1007/s10548-011-0205-y

60. Oh A, Duerden EG, Pang EW. The role of the insula in speech and language processing. Brain Lang. 2014;135:96–103. doi:10.1016/j.bandl.2014.06.003

61. Tetzloff KA, Duffy JR, Clark HM, et al. Longitudinal structural and molecular neuroimaging in agrammatic primary progressive aphasia. Brain. 2018;141(1):302–317. doi:10.1093/brain/awx293

62. Bocchetta M, Todd EG, Peakman G, et al. Differential early subcortical involvement in genetic FTD within the GENFI cohort. NeuroImage Clin. 2021;30:102646. doi:10.1016/j.nicl.2021.102646

63. McKenna MC, Chipika RH, Li Hi Shing S, et al. Infratentorial pathology in frontotemporal dementia: cerebellar grey and white matter alterations in FTD phenotypes. J Neurol. 2021;268(12):4687–4697. doi:10.1007/s00415-021-10575-w

64. Planche V, Mansencal B, Manjon JV, et al. Anatomical MRI staging of frontotemporal dementia variants. Alzheimers Dement. 2023;19(8):3283–3294. doi:10.1002/alz.12975

65. McKenna MC, Li Hi Shing S, Murad A, et al. Focal thalamus pathology in frontotemporal dementia: Phenotype-associated thalamic profiles. J Neurol Sci. 2022;436:120221. doi:10.1016/j.jns.2022.120221

66. McKenna MC, Hutchinson S, Bede P. The contribution of thalamic pathology to the clinical manifestations of frontotemporal dementia phenotypes (P5-12.004). In: *Monday, April 24*. Lippincott Williams & Wilkins; 2023:2956. doi:10.1212/WNL.0000000000202916

67. Janak PH, Tye KM. From circuits to behaviour in the amygdala. Nature. 2015;517(7534):284–292. doi:10.1038/nature14188

68. Woolley JD, Khan BK, Murthy NK, Miller BL, Rankin KP. The Diagnostic Challenge of Psychiatric Symptoms in Neurodegenerative Disease: Rates of and Risk Factors for Prior Psychiatric Diagnosis in Patients With Early Neurodegenerative Disease. J Clin Psychiatry. 2011;72(02):126–133. doi:10.4088/JCP.10m06382oli

69. Ducharme S, Dols A, Laforce R, et al. Recommendations to distinguish behavioural variant frontotemporal dementia from psychiatric disorders. Brain. 2020;143(6):1632–1650. doi:10.1093/brain/awaa018

70. Manera AL, Dadar M, Collins DL, Ducharme S. Ventricular features as reliable differentiators between bvFTD and other dementias. NeuroImage Clin. 2022;33:102947. doi:10.1016/j.nicl.2022.102947

